# Addressing medical and social needs to reduce unnecessary health care utilization and costs: a qualitative study

**DOI:** 10.1101/2021.11.11.21266203

**Authors:** David T. Liss, Adriana Guzman, Emily E. Walsh, Sara Shaunfield, Tiffany Brown

## Abstract

**Background:** There are few if any well-known approaches to reducing avoidable health care utilization and costs in patients with social needs. This study’s objective was to explore the goals, and approaches to organizing and delivering care, of interventions attempting to reduce avoidable resource use by addressing patients’ medical and social needs.

**Methods:** Semi-structured interviews were conducted with study investigators about early interventions in the peer-reviewed literature. A template analysis approach was used to review interview transcripts for common themes and create a final code list. Coder dyads separately coded each interview and resolved any discrepancies.

**Results:** Interviews were conducted with 15 investigators of interventions that delivered a variety of health services and addressed several individual social needs. Participants frequently described their overall goal as meeting patients’ diverse needs to prevent unnecessary acute care utilization. Reported approaches to addressing medical needs included assistance with receipt of primary care and care coordination across settings. Social needs were described as tightly linked with medical needs; addressing social support and housing were perceived as distinct from addressing other social needs. Participants described their overall approach to meeting patients’ needs in terms of establishing connections, partnership, respect, and being adaptable to patients’ priorities.

**Conclusions:** Findings shed new light on how to simultaneously address medical and social needs. Opportunities for future research include evaluating different approaches to addressing medical needs (primary care versus care coordination), separately evaluating the impacts of housing or social support, and hiring and training procedures to promote trauma-informed, patient-centered care.

## INTRODUCTION

The social determinants of health (SDOH) are a critically important driver of population health.^1, 2^ Recently, a growing understanding of the importance of SDOH— thrown into stark relief by inequities of the Covid-19 pandemic^3^—has driven a flurry of interest in addressing health-related social and economic needs (described henceforth as “social needs”) alongside patients’ medical needs in health care settings. For example, the Centers for Medicare & Medicaid Services is testing efforts to address beneficiaries’ housing instability, food insecurity, and other social needs,^4^ and leading private foundations have collaborated to identify evidence-based health care interventions for patients with social needs.^5^ In addition, many efforts to reduce avoidable health care utilization are being pursued in populations with immediate social needs, particularly in acute care settings such as hospitals. These efforts include federal initiatives to reduce inpatient readmissions,^6^ transitional care interventions,^7^ and programs to increase quality of life among high-risk patients living in community-based settings.^8, 9^

Nevertheless, there are few if any well-known approaches to reducing avoidable health care utilization and costs in patients with social needs. Organizations frequently face significant challenges when attempting to integrate social services with medical care,^10^ and a review of interventions to address social needs found mixed results on utilization or cost outcomes.^11^ Potentially promising observational findings are subject to multiple threats to internal validity—such as lack of a control group, and regression to the mean—as demonstrated by interventions such as the Camden Coalition of Healthcare’s hotspotting program, where cost reductions from initial pre-post observational data^12^ were not replicated in subsequent experimental research.^13^ Additionally, a variety of approaches may be needed to address the wide array of different social needs faced by patients; for example, interventions to resolve transportation barriers may bear little resemblance to those addressing housing instability or food insecurity.

There may thus be substantial value in examining and synthesizing early efforts to control health care utilization and costs by addressing patients’ medical and social needs. A summary of the goals, tactics, and insights of investigators in this area of inquiry can make important contributions to the evidence base by identifying common themes across interventions and potential new opportunities for investigation. The current study was conducted to explore the goals, and approaches to organization of delivery of care, across several interventions that sought to reduce avoidable resource use by simultaneously addressing patients’ medical and social needs.

## METHODS

### Study setting and participants

This qualitative study included semi-structured interviews conducted between November and December 2020. The study protocol was approved by Northwestern University’s institutional review board (IRB #STU00213468) and informed consent was obtained for all participants.

Potential participants were identified during an ongoing systematic review^14^ of published studies meeting three main inclusion criteria: 1) the study intervention addressed patients’ medical needs, with medical services delivered by a health care professional (or lay health worker); 2) the intervention addressed at least one of the following nine types of social needs: food, housing, interpersonal violence, legal services, social support, child care/adult day care, transportation, financial assistance, or inclusion of a social worker on the care team, and; 3) the study evaluated intervention impacts on utilization or costs of emergency department visits, inpatient admissions, or total health care costs. In addition, all eligible studies were conducted in non-institutionalized U.S. adults, and had a randomized trial or observational design with a control group.

For this qualitative study we recruited investigators of studies meeting the above inclusion criteria that had peer-reviewed results published in the year 2015 or later. During October and November 2020, we emailed corresponding authors of qualifying manuscripts to invite them to participate in a 45-to-60-minute interview via audio or video call. Some corresponding authors referred us to other project team members who could provide insights about the intervention. Potentially eligible investigators were contacted up to four times via email. Those who agreed to participate were scheduled for a one-hour call, and the verbal informed consent script was emailed to the participant along with scheduling details.

### Data collection and analysis

Semi-structured interviews were conducted by two study authors (DTL, TB), both of whom are health services researchers with experience evaluating quality improvement initiatives in primary care settings for vulnerable populations. In preparation for individual interviews, interviewers reviewed publications containing participants’ study results, which also enabled us to collect selected data on intervention characteristics (e.g., study design and geographic region). Interviews were recorded and auto-transcribed in Zoom (Zoom Video Communications, Inc.; San Jose, CA). Interviewers used a semi-structured interview guide that was designed to gain insight into the intervention’s goals, how the intervention addressed patients’ medical needs and social needs, investigators’ overall approach to supporting patients, and necessary skills and competencies for intervention team members (Table 1). At the end of each interview, the participant completed a brief survey about their demographic characteristics, training, and work experience.

**Table 1:** Semi-structured interview guide questions

| **Topic** | **Interview Guide Question** |
| --- | --- |
| Goals | What was the main goal, or the top 2 overarching goals, of the intervention for supporting patients? |
| Addressing medical needs | In what ways did your intervention address the medical needs of patients? |
| Addressing social needs | In what ways did your intervention address the social needs, or economic needs, of patients? |
| Overall approach to supporting patients | Now I'd like to take a step back to discuss the overall approach to supporting patients, if there was one. That is, above the level of any individual intervention components or individual patient needs, was there a larger overarching approach you took to achieve the goal(s) of the intervention? If so, could you describe it? |
| Staff skills/competencies | Can you tell me about any special training (i.e., clinical training, motivational interviewing, etc.), personal skills, or cultural competencies that were required for key members of the intervention team? |

Prior to data analysis, one team member listened to the audio from each interview, checked the recording against the auto-transcription, and made corrections when necessary. Relevant sections of each transcript were then transferred to Microsoft Excel (Microsoft; Redmond, WA) and formatted so responses to specific questions were in a single spreadsheet cell, creating a final matrix with one row for each participant and one column for each interview guide question.

Our coding approach employed template analysis,^15, 16^ where we initially created a priori codes to guide analysis and yield insights on targeted themes, and then refined the initial coding scheme to capture insights gleaned from the data. We began analysis by developing an initial code list centered around individual interview guide questions. Interviewers (DTL, TB) discussed common themes as interviews were still ongoing. After data collection, three coders (DTL, EEW, TB) met over several weeks to discuss contents of the transcripts, themes that emerged across all transcripts, and possible codes to add or collapse within the code list. We met again after each coder had coded two interviews on their own and, using a content analysis approach,^17^ expanded the initial code list to include additional codes. After every transcript was independently coded by two authors, differences were resolved via discussion within each coder dyad. Each participant response had a primary code and, if applicable, secondary codes.

## RESULTS

We recruited a total of 37 investigators from 31 different studies. During recruitment, one potential participant was deemed ineligible because they were an external evaluator who did not have intimate knowledge of intervention protocols. Five potential participants were not reached due to undeliverable email addresses, eight did not respond to recruitment emails, and seven declined participation. Sixteen participants completed interviews about 14 different interventions (two interviews included two investigators from the same study team). Following data collection, we excluded one completed interview from analysis because it examined a community-level intervention that did not target individual patients.

The final study sample included 15 investigators from 13 different studies (Table 2). Most participants (73%) were female, and 9 (60%) were physicians. The sample generally consisted of experienced researchers; 7 (47%) were their study’s principal investigator, and 8 (53%) worked at an academic medical center. Participants reported a mean of 17 years of research experience.

**Table 2:** Participant characteristics

| Characteristic | Participants (n=15)* |
| --- | --- |
| Female, n (%) | 11 (73%) |
| Degree, n (%) |  |
| MD | 9 (60%) |
| PhD/DrPH | 4 (27%) |
| Masters | 2 (13%) |
| Study PI, n (%) | 7 (47%) |
| Years in research, mean (SD) | 17 (9) |
| At academic medical center, n (%) | 8 (53%) |
|  | Interventions (n=13) |
| Study design, n (%) |  |
| Randomized controlled trial | 7 (54%) |
| Observational | 6 (46%) |
| Geographic region, n (%) |  |
| Northeast | 5 (38%) |
| Midwest | 1 (8%) |
| West | 3 (23%) |
| Southeast | 2 (15%) |
| Multiple† | 2 (15%) |

\* 2 of 13 interviews had 2 participants
† Intervention sites in more than one geographic region

There was substantial heterogeneity across the 13 studies. Seven (54%) were randomized trials, and study sites were geographically dispersed across the country. Studies evaluated medical interventions that delivered a variety of health services including transitional care, patient navigation, care coordination, and primary care with co-located wraparound services. Study interventions addressed individual social needs such as food insecurity, housing, social support/isolation, transportation barriers, and financial strain.

Several themes were identified from the qualitative data, which we report on here and include additional illustrative participant quotes in Table 3. When describing the main goal of their intervention, participants consistently mentioned a dual objective of simultaneously meeting patients’ needs while preventing unnecessary, high-cost forms of health care utilization (Table 3). Most commonly, participants stated that they aimed to have patients receive outpatient care as a substitute, or form of prevention, for downstream emergency department visits or inpatient admissions. Some participants explicitly stated that their intervention sought to achieve the Triple Aim of reduced health care costs, improved patient experience, and increased population health.^18^ Other participants described a broad goal of meeting previously unmet patient needs.

**Table 3:**
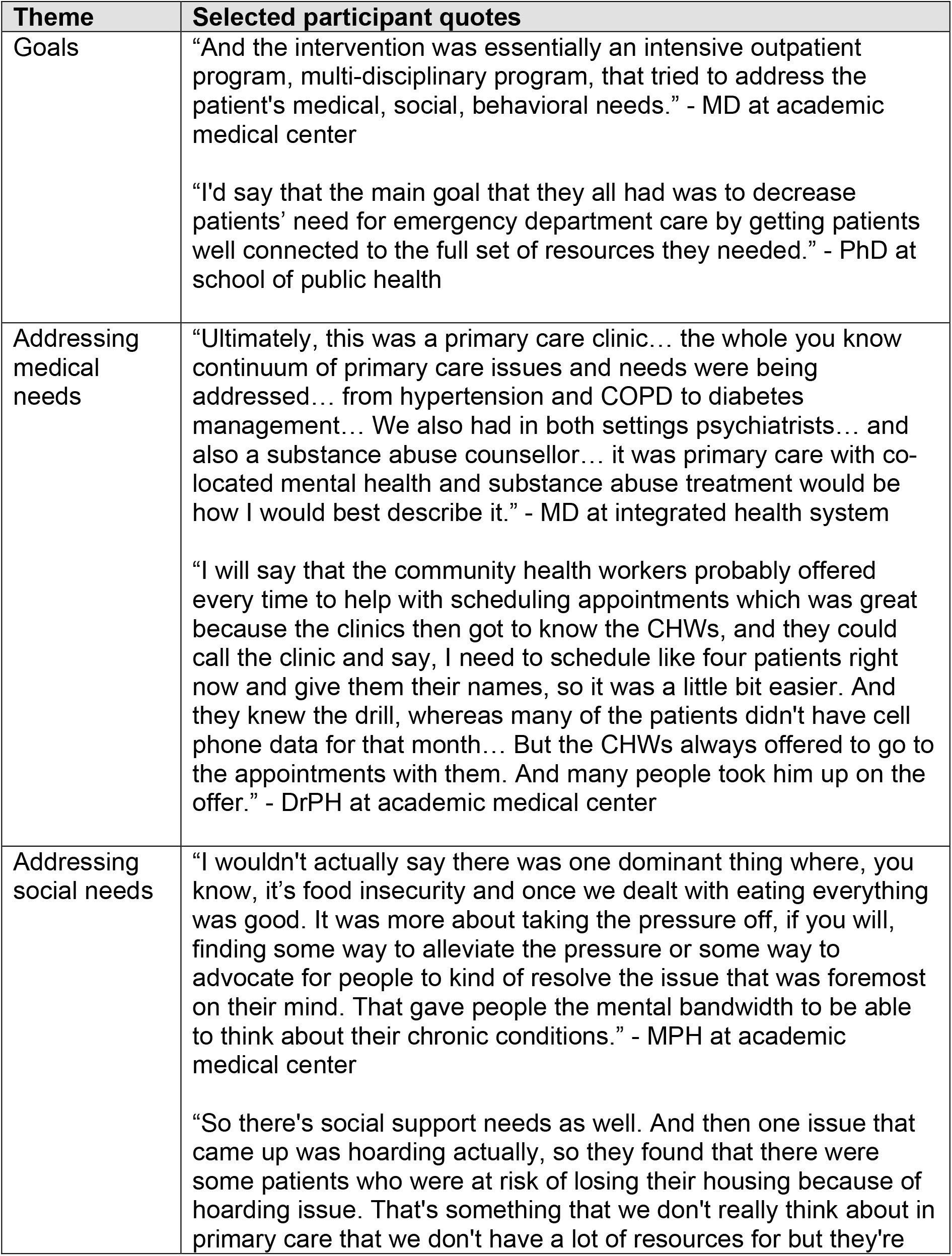
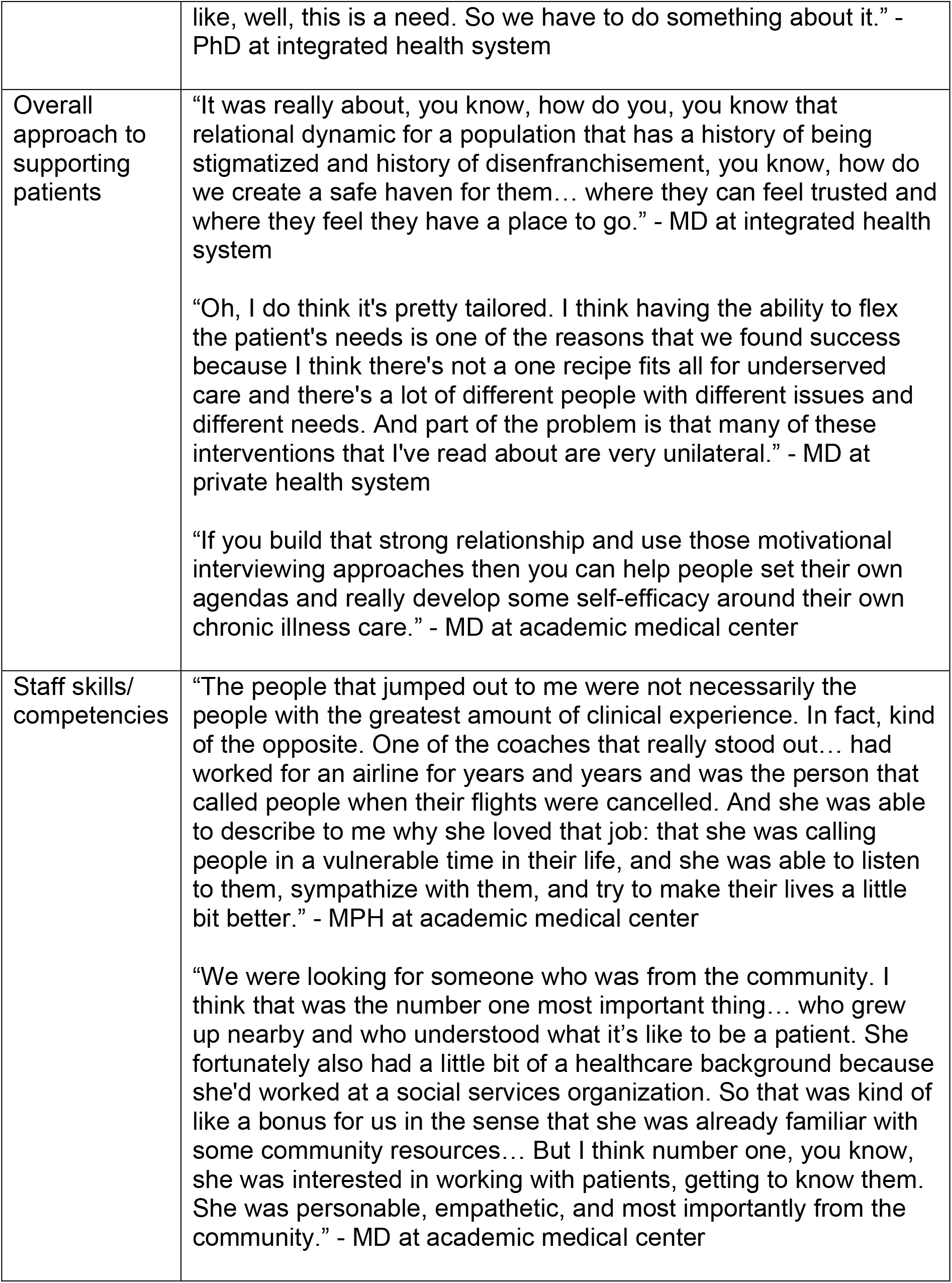
Themes and Illustrative Quotes

| Theme | Selected participant quotes |
| --- | --- |
| Goals | “And the intervention was essentially an intensive outpatient program, multi-disciplinary program, that tried to address the patient's medical, social, behavioral needs.” - MD at academic medical center<br>“I'd say that the main goal that they all had was to decrease patients' need for emergency department care by getting patients well connected to the full set of resources they needed.” - PhD at school of public health |
| Addressing medical needs | “Ultimately, this was a primary care clinic... the whole you know continuum of primary care issues and needs were being addressed... from hypertension and COPD to diabetes management... We also had in both settings psychiatrists... and also a substance abuse counsellor... it was primary care with co-located mental health and substance abuse treatment would be how I would best describe it.” - MD at integrated health system<br>“I will say that the community health workers probably offered every time to help with scheduling appointments which was great because the clinics then got to know the CHWs, and they could call the clinic and say, I need to schedule like four patients right now and give them their names, so it was a little bit easier. And they knew the drill, whereas many of the patients didn't have cell phone data for that month... But the CHWs always offered to go to the appointments with them. And many people took him up on the offer.” - DrPH at academic medical center |
| Addressing social needs | “I wouldn't actually say there was one dominant thing where, you know, it's food insecurity and once we dealt with eating everything was good. It was more about taking the pressure off, if you will, finding some way to alleviate the pressure or some way to advocate for people to kind of resolve the issue that was foremost on their mind. That gave people the mental bandwidth to be able to think about their chronic conditions.” - MPH at academic medical center<br>“So there's social support needs as well. And then one issue that came up was hoarding actually, so they found that there were some patients who were at risk of losing their housing because of hoarding issue. That's something that we don't really think about in primary care that we don't have a lot of resources for but they're |
|  | like, well, this is a need. So we have to do something about it.” - PhD at integrated health system |
| Overall approach to supporting patients | “It was really about, you know, how do you, you know that relational dynamic for a population that has a history of being stigmatized and history of disenfranchisement, you know, how do we create a safe haven for them... where they can feel trusted and where they feel they have a place to go.” - MD at integrated health system<br>“Oh, I do think it's pretty tailored. I think having the ability to flex the patient's needs is one of the reasons that we found success because I think there's not a one recipe fits all for underserved care and there's a lot of different people with different issues and different needs. And part of the problem is that many of these interventions that I've read about are very unilateral.” - MD at private health system<br>“If you build that strong relationship and use those motivational interviewing approaches then you can help people set their own agendas and really develop some self-efficacy around their own chronic illness care.” - MD at academic medical center |
| Staff skills/competencies | “The people that jumped out to me were not necessarily the people with the greatest amount of clinical experience. In fact, kind of the opposite. One of the coaches that really stood out... had worked for an airline for years and years and was the person that called people when their flights were cancelled. And she was able to describe to me why she loved that job: that she was calling people in a vulnerable time in their life, and she was able to listen to them, sympathize with them, and try to make their lives a little bit better.” - MPH at academic medical center<br>“We were looking for someone who was from the community. I think that was the number one most important thing... who grew up nearby and who understood what it's like to be a patient. She fortunately also had a little bit of a healthcare background because she'd worked at a social services organization. So that was kind of like a bonus for us in the sense that she was already familiar with some community resources... But I think number one, you know, she was interested in working with patients, getting to know them. She was personable, empathetic, and most importantly from the community.” - MD at academic medical center |

When describing the ways their intervention addressed patients’ medical needs, participants frequently mentioned one of two approaches. The most common approach was working to ensure patients received primary care, so primary care teams could identify and address comorbid medical conditions. Primary care sites often offered other co-located services, such as behavioral health and substance abuse treatment. In some instances, intervention staff co-attended primary care appointments with patients, which allowed them to do things like advocate on the patient’s behalf and become fully informed about treatment plans. Participants’ other frequently mentioned approach to addressing medical needs was active care coordination, in which intervention team members work to organize care that patients received, facilitate information transfer between stakeholders, and reduce fragmentation of care. Participants provided examples of how both approaches to addressing medical needs allowed them to achieve medication-related tasks, such as conducting medication reconciliation or working with clinicians to establish and streamline medication regimens that were feasible for patients.

Participants frequently conceptualized social needs as an important domain of need that was tightly linked with medical needs. Social needs were sometimes described as a barrier to medical care, or as pressing needs that “dramatically affect” how patients experience health care, which therefore had to be met before clinicians could effectively address patients’ medical needs.

When describing approaches used to address patients’ social needs, participants’ described similar approaches to the ways they addressed medical needs. Whereas primary care practices were often described as the principal setting for addressing medical needs, some participants stated that their intervention served as a central hub for identifying and addressing social needs. Additionally, intervention teams’ aforementioned care coordination tasks were not necessarily limited to addressing medical needs; some participants stated that their intervention served as a resource for identifying and navigating available community and social services.

Several individual types of social needs, such as food insecurity, transportation, and utility bill payments, were generally described as needs that were identified and then addressed (either directly through the intervention or by navigation to available resources). However, efforts to provide social support and address housing needs were perceived as distinctly different from efforts to address other social needs. For social support, several participants described efforts to create a welcoming, hospitable atmosphere at their practice—“how would you treat your grandma”—to make patients feel supported and increase their willingness to return for subsequent visits. Some practices offered creature comforts such as a television, food, showers, or new clothes. Creation of peer support groups was also described as a source of comfort and potential resilience for patients dealing with multiple stressors. Participants engaged in a variety of efforts to address housing-related needs, including referral to community resources for help paying utility bills and coordination of medical care at homeless shelters. Unfortunately, it was not always possible for participants to help patients obtain housing (e.g., a multiyear waitlist for local housing programs).

When describing their *overall* approach to meeting patients’ needs, participants frequently described a philosophy of connection and partnership, rooted in respect for patients’ experiences and values. A common theme was the need to take a flexible approach to meeting patients’ needs. Rather than directing the care planning process, participants mentioned the need to adapt to patients’ desires and priorities, so as to address modifiable factors that patients were ready to address. Additionally, participants frequently mentioned the need to build relationships with patients, and gain their trust, prior to then partnering to address previously intractable needs.

When identifying required skills and competencies for intervention team members, participants frequently mentioned having strong communication skills, empathy during interactions with patients, or membership in (or cultural humility towards) the local community. These skills were prioritized over explicit clinical training, though participants also stated that it was desirable for intervention staff to be generally familiar with health care systems.

## DISCUSSION

We observed several notable findings in this targeted qualitative investigation. Many participants stated that meeting patients’ multifaceted needs was their principal mechanism for preventing unnecessary, high-cost forms of utilization. Interventions most commonly worked to meet patients’ medical needs by facilitating receipt of appropriate primary care services or actively coordinating care across clinical settings. Many interventions served as the primary venue for directly addressing patients’ social needs, while others helped patients identify, navigate, and obtain available community and social services. Some participants perceived providing social support or addressing housing needs as distinct from efforts to address other social needs. Many participants described their overall approach to supporting patients in terms related to connection, partnership, and respect. Communication skills and empathy were frequently described as required competencies.

Several of our findings have parallels to results from prior studies. Similar to our finding that participants conducted care coordination to address patients’ medical needs, a recent evaluation of a home visiting program found that many high-need patients reported care coordination-related needs, such as making appointments with different types of providers and rescheduling missed appointments.^19^ Participants’ prioritization of communication skills, empathy, and knowledge of the local community align with staffing protocols of the IMPaCT community health worker program, which hired and rigorously trained “natural helpers” from the local community who had strong listening skills and were non-judgmental.^20^

Our findings also highlight the fact that complex patients with comorbid medical and social needs are often poorly served by traditional medical care settings. Participants’ responses highlight several of these overlapping factors, from the need to coordinate care across fragmented health systems to urgent social needs that have to be met before clinicians can effectively address medical needs. Many of the processes and principles described by participants align with those of trauma-informed care and patient-centered care. For example, just as participants identified the need for relationships with patients that are rooted in connection, partnership, and respect for patients’ prior experiences and values, trauma-informed care is characterized by partnering with clients, sharing of power, and promoting an environment where people feel physically and psychologically safe.^21^ Participants’ reported approaches to supporting patients, and involving them in health care decisions and delivery, are also in accordance with many aspects of patient-centered care.^22, 23^

Our findings point to several potential opportunities for research. Future studies could compare the two most common approaches participants used to address medical needs (i.e., comprehensive primary care versus care coordination across clinical settings). New research could also explore potential differences between the individual types of social needs that were identified by participants. For example, lack of housing or social support may be predictive of different adverse outcomes than other social needs such as food insecurity or transportation barriers. Additionally, programs that address different types of social needs may offer disparate—and potentially synergistic—benefits that could be identified within studies designed to evaluate multicomponent interventions.

This study has several limitations. Its sample size was small, and there was substantial heterogeneity across participants’ intervention settings. Some interventions were small, single-site programs, while others were multi-site programs across large health systems. All participants were investigators of interventions that aimed to control acute care utilization or costs, but not all of their evaluations demonstrated statistically significant reductions in these outcomes. Though a lack of success in this regard may have allowed participants to gain important insights about supporting patients and addressing their needs, it also means that some participants were describing interventions that did not necessarily achieve their aims. The research team’s health services research background could have affected the flow and content of interviews (e.g. which probes were asked), and our interpretation of results in the context of trauma-informed care and patient-centered care.

## Conclusion

In this qualitative exploration of interventions to reduce avoidable emergency and inpatient utilization and costs, participants reported meeting patients’ medical needs by facilitating primary care receipt or care coordination activities. Reported approaches to addressing social needs were somewhat similar. While overall approaches to supporting patients aligned with principles of trauma-informed care and patient-centered care, desired staff competencies focused on interpersonal skills. Study findings improve our understanding of interventions that simultaneously address medical and social needs, and point to many opportunities for future research.

## Supporting information

Checklist: Standards for Reporting Qualitative Research

Conflict of Interest Disclosure Form

## Data Availability

All data produced in the present study are available upon reasonable request to the authors.

## Acknowledgments

We thank Kenzie A. Cameron, PhD, Sara J. Doyle, MD, Lauren Leviton, LCSW, Matthew J. O’Brien, MD, and Caitlin A. Visek, MD, who helped establish study inclusion criteria and screened manuscripts to identify the sample of potentially eligible interview participants. Corinne Miller, MLIS supported development of the accompanying PubMed search, review of abstracts, and obtaining full text articles.

